# Longevity and frailty and their associated factors in the prognosis of coronary heart disease in a Latin American cohort

**DOI:** 10.1101/2023.10.16.23297091

**Authors:** Luis Andres Dulcey Sarmiento, Diego Andrey Acevedo Peña, Silvia Fernanda Castillo Goyeneche, Anderson Felipe Arias, María Juliana Estévez Gómez, Jaime Gomez, Carlos Hernandez, Juan Sebastián Theran Leon, Raimondo Caltagirone, Edgar Blanco, María Ciliberti, Silvia Yulieth Calderon Amaya, Emily Gutierrez, Angie Lizcano, Maria Camila Amaya, Diego Lobo, Francisco Mora Estupiñan

## Abstract

**Introduction:** to study the characteristics of the anamnestic history, the clinical course, as well as the nature of medical care for elderly patients with acute myocardial infarction (MI) and evaluate their impact on the prognosis of the disease.

**Material and methods:** the study included patients aged 60 years or older, survivors of acute myocardial infarction and registered in the database of the Hospital Universitario de los Andes in Merida, Venezuela (n=410). During the 2013-2018 study, a five-year prospective observation of patients with an assessment of their vital status was conducted. For the statistical processing of the data obtained, the Statistica V10.0 application package (StatSoft Inc.) was used.

**Results:** the analysis showed that 90% of the patients had a history of comorbid pathology. The presence of an atypical manifestation of myocardial infarction lengthened the prehospital stage of medical care for patients who were late in seeking medical attention (120 [49; 311.5] minutes), as well as the longest time until the first medical contact (26.5 [20; 40] minutes (p = 0.005)). One-fifth of patients were treated for acute MI in complementary hospitals, where the level of in-hospital mortality among elderly patients reached 65.7%, three times higher than the same figure in specialized cardiology departments (19, 7%, p< 0.001).

**Conclusion:** The main factors influencing long-term post-infarction in elderly patients were isolated systolic arterial hypertension, diabetes mellitus, renal dysfunction, previous myocardial infarction and acute stroke.

## INTRODUCTION

Acute myocardial infarction (MI), as is known, is one of the leading causes of death and disability in older age groups [1]. For various reasons, caused by a complex of physiological changes that develop during the aging process, and pathological processes that arise as a result of the accumulation of polymorbidity, patients in this age group have certain characteristics in terms of clinical picture, diagnosis, treatment of acute myocardial infarction and the course of the post-infarction period. According to the literature, the vast majority of people develop polymorbidity at the age of 60 years; after the age of 75, the number of combined diseases is usually 4 to 6. Less than 7% of older people are practically healthy and do not suffer from serious chronic diseases [2]. Along with this, in the presence of concomitant pathology, mortality at three years increases progressively and exceeds 80% in the presence of two or more concomitant diseases [3].

At the same time, the combination and overlap of various pathophysiological mechanisms for the development of the main and concomitant diseases, the constant use of medications, often forced polypharmacy and a decrease in the reactivity of the organism determine an increased frequency of erased and atypical diseases. forms of acute myocardial infarction [4] [5]. It should also be taken into account that elderly and senile people often suffer from various psychoneurological disorders, accompanied by a distortion of patients’ perception of the symptoms of coronary heart disease, which leads to an underestimation of its severity [6] [7].

Therefore, the diagnosis and treatment of MI in elderly patients may present certain difficulties. Taking into account that the epidemiological situation of coronary heart disease and, in particular, acute myocardial infarction, is determined by the frequency of its development mainly among people aged 60 years and older, the study of this topic is relevant and appropriate.

The aim of the study was to study the characteristics of the anamnestic history, the clinical course, as well as the nature of medical care for elderly patients with acute myocardial infarction and evaluate their impact on the prognosis of the disease.

## MATERIAL AND METHODS

The study included patients aged 60 years or older, survivors of acute myocardial infarction and registered in the database of the Hospital Universitario de los Andes (n=410). During the study from 2013-2018, a five-year prospective observation of the patients with an assessment of their vital status was carried out. The study protocol was approved by the Biomedical Ethics Committee of the Research Institute where it was carried out.

Statistica V10.0 application package (StatSoft Inc.) was used. The normality of the distribution of the parameters was checked using the Shapiro-Wilk test. Taking into account the non-compliance with the normal law distribution, the average values of the quantitative data are presented as median and interquartile range (Me (Q25; Q75)). Comparisons in two independent samples were performed using the Mann-Whitney U test. Qualitative data are presented in the form of absolute and relative values, the significance of differences of which was assessed using the Pearson test (χ2) as well as a two-sided Fisher’s exact test. When testing hypotheses, the critical level of statistical significance was set at p ≤ 0.05. To analyze survival, as well as the time until the appearance of adverse cardiovascular events, the Kaplan-Meier method was used to construct the corresponding curves. A comparative analysis of the graphs obtained was performed using the log-rank test. To identify the factors most associated with adverse events and disease prognosis, odds ratios (OR) were calculated.

## RESULTS

The mean age of the 410 patients included in the study was 71 [66; 77] years (for men - 69 [63; 74] years, for women - 74 [69; 80] years, p < 0.001), 51.7% of participants were men. A combination of three or more risk factors (RF) for cardiovascular complications occurred in 90% (n=369) of patients. Furthermore, in the vast majority of cases, patients in the study group had a history of comorbid pathology. In a five-year prospective follow-up, the mortality rate reached 42.4% (n=174).

High blood pressure (AH) occurred in almost 90% of patients, and more than half of them had isolated systolic high blood pressure (HAIS), which is known to be associated with the risk of adverse cardiovascular events and mortality, which was confirmed by in our study. Thus, ISAH was recorded more frequently in the group of patients who died within five years of follow-up after the index event (n=174), increasing the probability of developing an unfavorable outcome by more than 1.5 times (64.1% versus 51.7 %, OR - 1.7; 95% CI: 1.09–2.56; p=0.016).

20% of patients had type 2 diabetes mellitus. The presence of this disease caused a nearly twofold increase in the five-year mortality rate in the study cohort (22% versus 33.9%; OR - 1. 82; 95% CI 1.17-2.82; p = 0.002).

In 33% of cases, the index MI recurred in the patients, another 27% suffered from exertional angina. It should be noted that in 40% of cases, patients in the study cohort did not seek medical help for coronary heart disease before the development of MI. 13% of patients (n=53) had a history of acute cerebrovascular accident (ACVA). At the same time, both MI and stroke, independently of each other, caused a significant increase in five-year mortality (OR - 2.2; 95% CI 1.43–3.3; p < 0.001; OR - 3.05; 95% CI 1.66– 5.59; p<0.001, respectively).

16% of patients had stage C3a chronic kidney disease (CKD); More than half of the patients (56%) showed a decrease in GFR to 60-89 ml/min/1.73 m2, which corresponds to stage C2 of CKD and taking into account the presence of markers of kidney damage. such as albuminuria, electrolyte alterations, etc., is a risk factor for the development of complications and can worsen both the immediate and long-term prognosis of the disease [8], as confirmed by our data. Furthermore, according to the results obtained in this study, the long-term prognosis of the disease was also negatively affected by a slight decrease in GFR in the range of 60 to 89 ml/min./1.73 m2 (OR - 5. 9; 95% CI 2, 9-11.9, p<0.001). At the same time, the prognosis of the disease worsened in proportion to the decrease in GFR, and this factor, in statistical analysis, was also shown to be an independent predictor of an unfavorable outcome in a five-year follow-up.

It should be noted that risk factors such as smoking were confirmed by 29% (n=118) of the patients, and the vast majority of them were men. Just under half of the patients (43%) were overweight and another 29% were obese. At the time of the development of an acute coronary accident, the majority of patients (84%) already had confirmed alterations in the composition of blood lipids. The development of index MI in the context of existing CHF occurred in 69% of patients (n=278) (Table 1).

**Table 1.** Anamnestic history in elderly and senile patients who have suffered acute myocardial infarction.

| **Variable** | **Result** |
| --- | --- |
| Smoking, n (%) | 118 (28.8) |
| Arterial hypertension, n (%) | 357 (87) |
| Isolated systolic arterial hypertension, n (%) | 204 (49.8) |
| Diabetes mellitus type 2, n (%) | 111 (27.1) |
| Obesity, n (%) | 117 (28.5) |
| Excess body weight, n (%) | 176 (42.9) |
| Myocardial infarction, n (%) | 134 (32.7) |
| Angina pectoris, n (%) | 245 (59.8) |
| Chronic heart failure, n (%) | 278 (67.8) |
| Stroke, n (%) | 53 (12.9) |
| Chronic obstructive pulmonary disease, n (%) | 60 (14.6) |
| Chronic kidney disease, n (%) | 45 (11) |
| Glomerular filtration rate, ml/min/1.73 m<sup>2</sup> · Me [Q25; P75] | 80 [62; 89] |
| Low-density lipoproteins, mmol /l | 3.3 (2.9; 4.0) |
| Glucose, mmol /l | 5.7 (5.3; 6.5) |
| Note: I [P25; Q75] – median and interquartile range . |  |

When analyzing the clinical picture of acute MI, it was found that one in five patients in the study group (17.8%) had an atypical clinical picture of the disease. Patients with an atypical course of the disease were on average 3 years older than patients with a classic angina attack (74 [66.5; 79] vs. 71 [64.5; 77], p = 0.029).

One in five cases in elderly and senile people was characterized by an atypical course of acute myocardial infarction. Furthermore, in 41% of atypical cases of manifestation of a coronary catastrophe, the clinical picture was represented by a poorly symptomatic form, which was more common in men (51.2% versus 28.1%, p = 0.037).

An interesting fact is that patients with an atypical clinical picture of the disease sought medical help within an average of two hours (120 [49; 311.5] minutes). Furthermore, 10% of elderly patients sought medical help more than 12 hours after the first symptoms of acute myocardial infarction appeared. At the same time, the time from the moment of request to contact with a medical professional was 26.5 [20 ;40] minutes (vs. 18 [16;29] minutes with a typical clinical picture; p = 0.005)). Overall, hospitalization time for patients in the study cohort averaged 4 hours (240 [150, 450] minutes).

One fifth of the patients in the study cohort (19%) were hospitalized with acute myocardial infarction in non-specialized general medical and surgical hospitals without the possibility of providing high-tech medical care. These facts seem to contribute greatly to the very high in-hospital mortality rates due to acute myocardial infarction among elderly patients treated in secondary hospitals, which reached 65.7%, three times higher than the same figure in specialized departments (19.7%)., p<0.001).

## DISCUSSION

Thus, most patients of older age groups suffer from MI against the background of pre-existing concomitant diseases, the presence of which, even individually, leads to a worse prognosis and reduces long-term survival rates, as confirmed by the literature. data [9] [10] [11]. However, along with this, the presence of polymorbidity itself, due to forced polypharmacy, may contribute to an increase in the number of atypical variants of the course of acute MI. Thus, according to large studies (Global Registry of Acute Coronary Events; National Registry of Myocardial Infarction), the frequency of cases of myocardial infarction, which occur with erased clinical symptoms, progressively increases with age, reaching 60% in older people 85 years old.. Furthermore, in more than 30% of cases of atypical clinical symptoms, the collaptoid variant occurs and in another 20%, the asthmatic variant occurs [5▪]. In our study, the most common atypical manifestation of acute MI was the poorly symptomatic variant. The lack of specificity of the clinical symptoms of acute MI in its atypical forms may cause patients to seek medical help late. Furthermore, the absence of classic manifestations of a coronary accident makes the diagnosis of myocardial infarction difficult in the prehospital stage, also increasing hospitalization time and increasing mortality rates [12▪]. For the same reason, patients with unexpressed and “non-classical” symptoms in 20% of cases are hospitalized in general medical and surgical hospitals, which, undoubtedly, cannot but affect the prognosis of the disease, taking into account the lack of possibility of timely myocardial revascularization [13].

The data presented indicate the need for increased attention to patients of older age groups, improvement of measures related to clinical examination to identify, timely correction of risk factors, treatment of concomitant pathologies and secondary prevention of cardiovascular events, especially considering that in 40% of patients before the development of the MI index, a relationship with coronary heart disease had never been observed. The education of patients in this cohort is also of great importance, since it has two objectives: on the one hand, raising patient awareness increases adherence to treatment and improves the effectiveness of secondary prevention; On the other hand, it teaches patients to pay attention to important changes in well-being and seek medical help in a timely manner [14]. At the same time, attention by medical workers to atypical, including asymptomatic, forms of acute myocardial infarction in people of this age category can help improve survival rates by timely sending patients to institutions specialized doctors.

## CONCLUSION

Thus, patients from older age groups with acute MI are characterized by the presence of a history loaded with comorbid pathology, the frequent presence of an atypical clinical picture of the disease, a high frequency of hospitalizations in complementary hospitals and high hospitalization rates. mortality. The main factors influencing the long-term post-infarction period in this patient cohort were the presence of ISAH, diabetes, previous MI and stroke, and renal dysfunction.

## Data Availability

All data produced in the present work are contained in the manuscript

